# Comparing Patient Reported Outcomes to Objective Measures of Function in patients with Chronic Pain: Using the SF-12, 2 minute walk test and Elevation and Movement Lift test

**DOI:** 10.1101/2021.05.07.21256342

**Authors:** Gaurav Gupta, Emilie Paquet-Proulx, LCol Markus Besemann, Kira Burton, Sasha Lalonde, Amir Minerbi

## Abstract

**Introduction:** An Ideal battery of testing for function would be inexpensive, easily administered, standardized and validated for multiple health issues. This would also be sensitive to change over time and able to extrapolate avocational and vocational tasks. The data collection exercise for this study included both subjective and objective measures which include the Short Form Health Survey 12 (SF-12), the 2 minute walk test (2MWT) and a newly developed upper extremity strength/conditioning activity called the Elevation and Movement Lift test (EMLi).

**Methods:** A convenience sample of 102 patients with chronic pain. They were seen in the Canadian Forces Health Services Unit (CFHSU (O)) Physiatry Clinic between January-September 2019 and were asked to complete the data intake protocol. This included: completing a questionnaire with the Numeric Rating Pain Scale (NRS) covering the previous 7 days, the SF-12, and completed the 2 MWT and EMLi.

**Results:** For the 2MWT heart rate, perceived exertion and number of steps were all increased for patients with chronic pain compared to the control group. There was no difference noted between patients with upper/lower body pain. In patients with chronic pain SF-12 physical function score negatively correlated with perceived exertion but not performance. As for the EMLI test, all groups had similar perceived exertion and heart rate outcomes but a reduced performance was noted with the upper extremity group.

**Discussion:** For the 2MWT, the individual’s performance related to effort and not their pain state, PE and SF-12. This suggests a higher capacity for walking then the patients realise. As for the EMLi, individual’s performance was poorer for same level of effort. This correlates to their perceived function as seen on the SF-12 which might measure pain related dysfunction.

**Conclusion:** The 2MWT performance was effort dependent and not correlated with perceived abilities. Therefore it can be used to challenge patient performance. EMLi performance correlated with perception and upper extremity pain. This could be used to set clinical training targets and monitor each individual’s progress.

## Introduction

The current assessment of functional status in patients with chronic pain relies primarily on patient reported outcomes in the absence of objective standard measures of physical function. Since patients with chronic pain have a complex interplay of non-reversible health issues, medications and psychosocial issues, having a clear understanding of actual function is of paramount importance in pain reduction targets and activity. Ideally, a battery of such tests for this purpose would be inexpensive, easily administered, standardized and validated for multiple health issues; while also being sensitive to change over time and able to extrapolate to avocational and vocational tasks.

When designing such a data collection exercise, both subjective and objective measurements should be used in conjunction. In one study, patients with chronic pain were; older, more likely to be overweight/obese, had lower mental health scores, lower 6-min walk results, sit-to-stand and stair-climbing performance in comparison to healthy controls. The authors emphasized assessing pain-related lifestyle risk factors and disability in addition to prevalence when considering the disease burden of pain [1]. In another study patients with fibromyalgia (FM) had worse perception of function on the SF-36, than actual physical function on the Senior’s Fitness test, and higher catastrophizing was consistently associated with this relationship. The authors noted that high catastrophizing may promote a feeling of reduced ability to do meaningful, daily activities than people with FM are actually able to do [2].

While numerous patient reported functional outcomes exist (i.e. 12 item Short Health Survey SF −12), there is dearth of standardized physical tests for patients with chronic pain [3,4]. Some of the current tests included the Senior Fitness Test battery [5], repeated sit-to-stand test (RSST) and daily pedometry [6]. Measuring distance walked in a given time period (i.e. 6 (6MWT) and 12 (12MWT) minute walk test) has been studied in healthy populations, patients with cardiopulmonary disease, and patients with chronic pain, and has been shown to have good validity and interrater reliability depending on the group studied [4,7,8,9,10,11,12,13].

This approach also allows for comparison of population norms when interpreting outcomes. One study showed the average distance walked for patients with chronic pain, averaging 40 years old, improved from 272.87 to 339.04 yards at 3-6 months, but still remained below the expected threshold for healthy adults of age 60 years [4]. The same authors suggested improving sensitivity with multiple tests including “a test for upper body strength” which as of yet has not been developed [4].

One strategy consists of using the SF-12, the 2 minute walk test (2MWT) and an upper extremity strength/conditioning activity. The short form health survey is an instrument for measuring health perception in a general population [14,15]. It has been used in patients with stroke [16], osteoarthritis [17] and rheumatologic [18] conditions as well as the elderly. The 2MWT has been either validated or used as a measure of function in patients’ with osteoarthritis [19], stroke [20] and amputation [21]. While numerous patient reported outcomes such as the Upper Limb Functional Index, Disabilities of the Arm, Shoulder and Hand (DASH) Questionnaire and Patient-Specific Functional Scale [22,23] have been proposed, we could not identify any physical measures similar to the 2 minute walk test. Therefore, we created a new physical measure for upper extremity function and conditioning known as the Elevation & Movement Lift test (EMLi).

The Canadian Armed Forces (CAF) has limited data on baseline quality of life measures and objective measures of function, for active serving members with chronic pain. This study aims to collect this data using SF-12 and 2MWT and newly created EMLi.

Our study proposes the following advantages and hypothesis:

1. Unique to the CAF members living with chronic pain;
2. Document point prevalence of severity of chronic pain issues within the CAF;
3. Patient reported health measures will be poorer than the control group;
4. Objectively, patients will have poorer performance for 2MWT compared to the control group;
5. Objectively, patients will have poorer performance for the EMLi when compared the control group;
6. Patients with neck pain and upper limb pain disorders will have poorer scores on the EMLi compared to the 2MWT;
7. Patients with low back pain and lower limb pain disorders will have poorer scores on the 2MWT compared to the EMLi.

## Methods

A convenience sample of 102 patients with chronic pain seen in the Physiatry clinic between January and September 2019 were asked to complete the data intake protocol. The CAF Health Surgeon General, provided approval for the data collection as a clinical quality improvement project.

The data intake protocol included completing a questionnaire with the:

1. Numeric rating pain scale (NRS) from 0-10 over the previous 7 days (least, average and greatest);
2. Short-Form Health Survey (SF-12);
3. Body Pain Diagram;

The physical activity measures taken were:

1. 2 minute walk test (2MWT);
2. Elevation & Movement Lift test (EMLi).

For both physical activity measures, patients’ heart rate was measured pre and post activity along with their response for the Borg perceived exertion scale. Between tests the patients’ heart rate was allowed to return to baseline, but for convenience and consistency, the 2MWT always preceded the EMLi.

The 2MWT was conducted in an unobstructed hallway with patients wearing the same electronic activity monitor (GARMIN vivofit4). Patients were asked to walk at moderate intensity (brisk walk) for the duration of the activity (2 minutes). The amount of steps were recorded from the activity tracker.

The EMLi movement consists of pushing a 2kg weighted ball from the chest level forward, and then raising the ball overhead, before returning it to the starting position (Figure 1). The number of repetitions done at moderate intensity was recorded after 45 seconds. The EMLi was first validated in 20 healthy controls before used in patients so parameters around weight and time for testing could be established.

**Figure 1:**
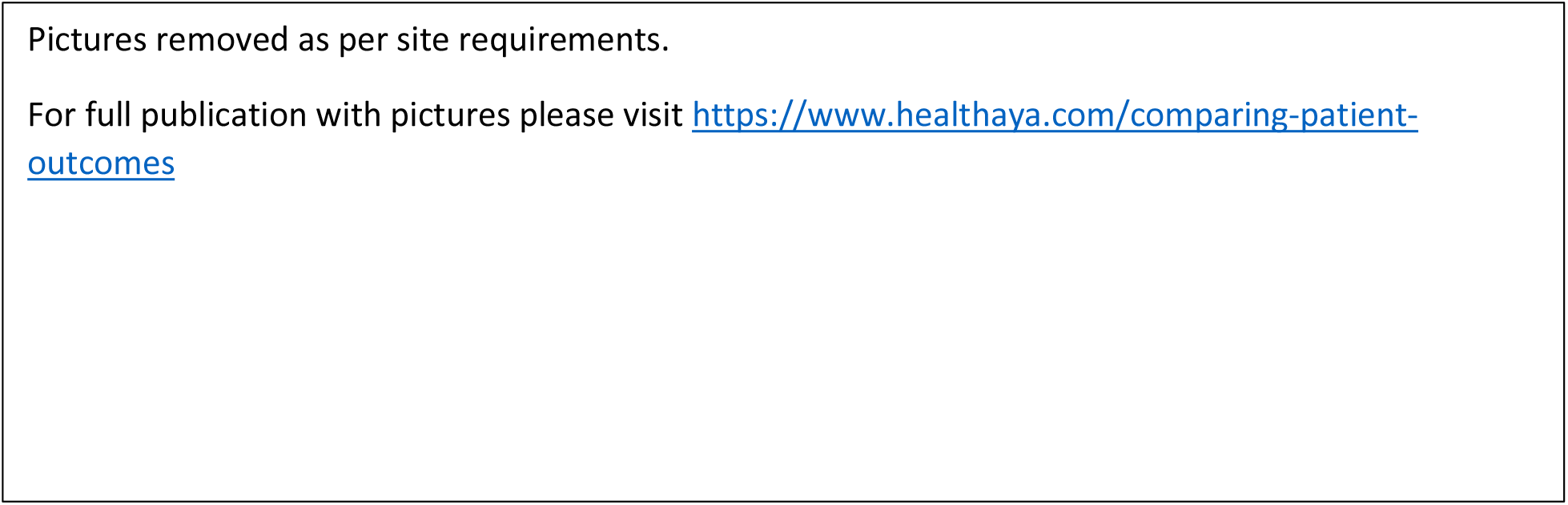
A demonstration of the EMLi test where a 2kg weight ball is pushed from the chest level forward (A to B), then raised overhead (C to D), before returning it to the starting position (A).

### Statistical analysis

The difference in performance during the upper and lower repetition tests were evaluated between the groups. This included heart rates, number of repetitions and perceived exertion. They were evaluated using multiple ANOVA (analysis of variance) tests, corrected for multiple comparisons using the Games-Howell post-hoc test.

Correlation analysis between selected variables was done using multiple Pearson correlations, and adjusted for multiple comparisons using the Benjamini-Hochberg false discovery rate equation.

## Results

When comparing the number of exercise repetitions and perceived exertion there were significant differences observed between chronic pain patients and healthy controls. Patients with either upper- or lower-body pain walked more steps (ie more repetitions) in the 2-minute walk test as compared to healthy controls. While healthy controls and patients with lower-body pain were able to perform better in the 45 second upper body repetition exercise (EMLi) than the patients with upper-body pain. Patients with either upper-or lower-body pain reported significantly higher perceived exertion following the 2-minute walk test, but not following the 45 second EMLi test. Finally, both pain patient groups experienced significantly higher heart rate increase following the 2-minute walk test but not following the 45 second EMLi test (Figures 2 and 3).

**Figure 2:**
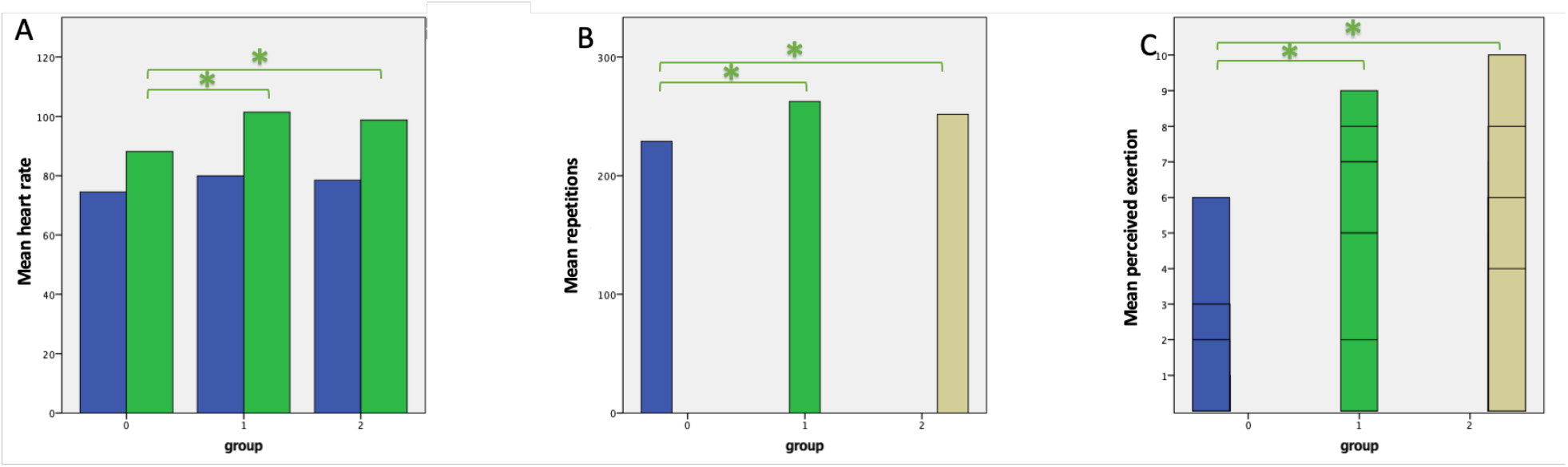
Between-group comparison of **(A)** mean heart rates before and after exertion; **(B)** mean number of repetitions achieved and **(C)** mean perceived exertion following the two-minute walk test. Group 0 – healthy controls; group 1 – upper-body pain patients; group 2 – lower-body pain patients. * Games-Howell post hoc test, p<0.05.

**Figure 3:**
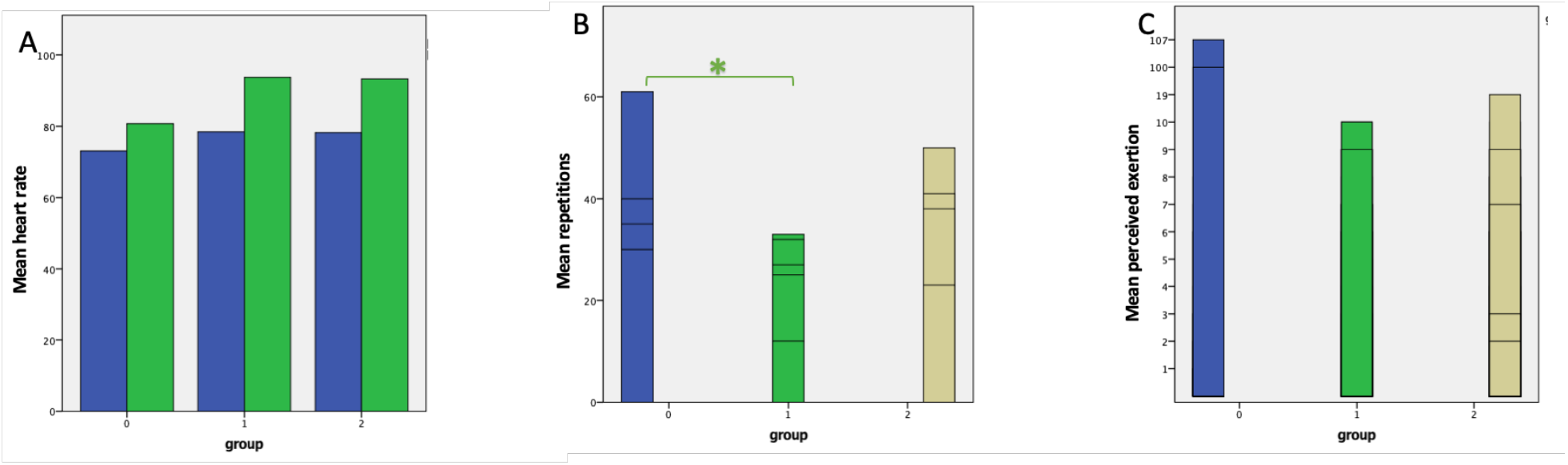
Between group comparison of **(A)** mean heart rates before and after exertion; **(B)** mean number of repetitions achieved and **(C)** mean perceived exertion following the 45 second upper body repetition test. Group 0 – healthy controls; group 1 – upper-body pain patients; group 2 – lower-body pain patients. * Games-Howell post hoc test, p<0.05.

The correlation between the different measured variables was explored using Pearson’s correlation corrected for multiple comparisons. SF-12 physical function score negatively correlated with perceived exertion but not performance with no impact correlation with Mental Health sub score.

## Discussion

This study represents the first of its kind within the Canadian Armed Forces correlating patient reported outcomes with physical capabilities while developing a new physical activity measure for looking at upper extremity function.

For the 2MWT, heart rate, perceived exertion and number of steps were all increased for patients with chronic pain compared to the control group. There was no difference between patients with upper and lower body pain. The SF-12 physical function score negatively correlated with perceived exertion and not performance. This indicates that patients with chronic pain may be able to do more physical activity that they believe that they can.

For the EMLi all groups worked at about the same perceived exertion and heart rate, but we still noted a difference in the upper extremity group (reduced performance). This indicates that this tool might be useful in a clinical setting to assign severity of injury and monitor clinical improvement over time.

There contrasts with previous work. One study found that women diagnosed with FM during the 6MWT had decreased levels of functional abilities and increased exacerbation of pain and exertion in comparison to healthy controls [24]. In a previous study, fibromyalgia (FM) patients and healthy controls self-reported similar physical activity levels. However, the FM patients had decreased physical performance on the 6MWT when compared to the control group. The data showed that women with FM actually perform about the same amount of physical activity as women without FM but exhibit decreased physical performance possible due to the lifestyle generally adopted by FM patients [25]. Perhaps perception of function and measured abilities in patients with chronic widespread pain are better correlated than the regional pain disorders seen in our studies.

While this study did not look longitudinally, one study did show improvement in 6MWT results from intake to follow up in patients with chronic pain at a single site [4]. In another study, participants with chronic pain had reduced capacity for physical activity. The pedometry results illustrated a range of maladaptive strategies adopted by those with chronic pain. From these results the authors suggested a subgroup of patients with chronic pain appear to avoid physical activity, potentially leading to greater disability and possibly deconditioning. Another subgroup appeared to persevere despite their pain leading to further pain and flares. Both behavioural responses may result in maladaptive biomechanical and central nervous system effects The method used to target physical activity in these patients should be considered in treatment planning, specifically for physiotherapy [26].

Limitations of this study include the relatively small sample size, the heterogeneity of chronic pain conditions, and the inability to control for various comorbid health conditions. In addition, measured distances instead of number of steps walked should be evaluated to avoid introducing a stride length bias. In the future, more strict performance examples by testers would be favorable.

Future studies can look to validate the EMLi in various other populations including healthy sub groups, patients with various spinal pain and localized joint dysfunction/musculoskeletal issues and cardiopulmonary conditions. Ideally, specific parameters such as the amount of weight required, timing of trials, reproducibility, validity and inter/intra-rater reliability need to be established as well. Following patient’s over time and linking this specific task could also be useful in developing clinical prediction rules regarding suitability for specific avocational and vocational activities.

## Conclusions

2MWT performance was effort dependent and not correlated with perceived abilities. Therefore this data can be used to challenge patient performance. EMLi performance correlated with perception and upper extremity pain. Therefore this data could be used to set clinical training targets and monitor progress.

## Data Availability

the data is available.

https://www.healthaya.com/comparing-patient-outcomes

